# Schizophrenia-associated somatic copy number variants from 12,834 cases reveal contribution to risk and recurrent, isoform-specific *NRXN1* disruptions

**DOI:** 10.1101/2021.12.24.21268385

**Authors:** Eduardo A. Maury, Maxwell A. Sherman, Giulio Genovese, Thomas G. Gilgenast, Prashanth Rajarajan, Erin Flaherty, Schahram Akbarian, Andrew Chess, Steven A. McCarroll, Po-Ru Loh, Jennifer E. Phillips-Cremins, Kristen J. Brennand, James T. R. Walters, Michael O’ Donovan, Patrick Sullivan, Psychiatric Genomic Consortium Schizophrenia and CNV workgroup, Brain Somatic Mosaicism Network, Jonathan Sebat, Eunjung A. Lee, Christopher A. Walsh

## Abstract

While inherited and *de novo* copy number variants (CNV) have been implicated in the genetic architecture of schizophrenia (SCZ), the contribution of somatic CNVs (sCNVs), present in some but not all cells of the body, remains unknown. Here we explore the role of sCNVs in SCZ by analyzing blood-derived genotype arrays from 12,834 SCZ cases and 11,648 controls. sCNVs were more common in cases (0.91%) than in controls (0.51%, p = 2.68e-4). We observed recurrent somatic deletions of exons 1-5 of the *NRXN1* gene in 5 SCZ cases. Allele-specific Hi-C maps revealed ectopic, allele-specific loops forming between a potential novel cryptic promoter and non-coding cis regulatory elements upon deletions in the 5’ region of *NRXN1*. We also observed recurrent intragenic deletions of *ABCB11*, a gene associated with anti-psychotic response, in 5 treatment-resistant SCZ cases. Taken together our results indicate an important role of sCNVs to SCZ risk and treatment-responsiveness.

## Introduction

*De novo* and rare germline CNV (gCNVs) contribute to up to 5.1-5.5% of SCZ cases, with relatively large effect sizes (Kirov et al., 2012). These gCNVs are usually inherited or, in the case of *de novo*, are thought to arise during gametogenesis. Most of gCNV involve several genes making it difficult to pinpoint specific causative genes. A notable exception is deletion of the *NRXN1* gene, which encodes a presynaptic adhesion protein, and has been suggested to have a role in SCZ along with other synaptic genes (Kirov et al., 2009).

Somatic CNV (sCNV), present in only a fraction of cells in the body, often represent mutations that are challenging to study in the germline state due to embryonic lethality or severe phenotypic impacts, and are increasingly implicated in neuropsychiatric disease. A recent study (Sherman et al., 2021) showed enrichment of large (>4 Mb) sCNVs in Autism Spectrum Disorder (ASD), with sCNV size positively correlated with phenotypic severity. The overlap in the genetic architecture of ASD and SCZ (Kushima et al., 2018) suggests that sCNV may have similar role in SCZ liability.

sCNV are less common than germline gCNVs, so that large datasets need to be analyzed to capture their contribution to disease. Whereas the largest genotyping datasets come from blood-derived SNP array data created for GWAS studies, assessing sCNVs in blood has been difficult because aging and environmental exposures such as smoking in SCZ patients create clonal hematopoiesis (CHIP) events as confounders. A previous study of blood derived SNP-array data from 3,518 SCZ cases and 4,238 controls showed a nominal increase of sCNVs in SCZ, but did not specifically detect mosaic events, or filter CHIP events, and defined sCNVs as larger than 10 Mb, limiting their characterization (Ruderfer et al., 2013).

In this study we analyzed SNP-array data from 12,834 cases and 11,648 control from the Psychiatric Genomic Consortium (PGC) SCZ cohort using a recently developed, highly sensitive algorithm that leverages haplotype information to detect sCNVs (Loh et al., 2018, 2020; Sherman et al., 2021), and rigorously filtered candidate variants that likely originated from CHIP, which have now been extensively characterized in multiple studies in terms of size, mosaic fraction and chromosomal location (Loh et al., 2018, 2020; Terao et al., 2020). We observed a robust excess of sCNVs in SCZ compared to controls, and discovered recurrent sCNVs with likely causative roles, including recurrent *NRXN1* somatic deletions of exons1-5. Taken together these data suggest a potentially important role of sCNVs in the genetic architecture of SCZ.

## Results

### Somatic CNVs are more prevalent in Schizophrenia cases than controls

Somatic CNVs were identified using the MoChA (Loh et al., 2018) pipeline on 26,186 blood-derived SNP arrays from the PGC2 SCZ cohort (Marshall et al., 2017). This pipeline exploits long-range haplotype-phasing information to detect sCNVs with high sensitivity (Loh et al., 2018, 2020). We used gCNVs previously identified in the subjects of this cohort (Marshall et al., 2017) to filter out potentially misclassified variants. Samples that showed signs of contamination or sCNVs whose copy number state was not confidently determined were excluded (Methods).

We employed a conservative filtering strategy to remove 1,032 events that could have risen from CHIP, as these events might bias burden estimates (Loh et al., 2018, 2020). Namely, we removed all copy-neutral loss of heterozygosity (CN-LOH), loci commonly altered in the immune system (e.g. Major Histocompatibility Locus (MHC)), and other known common CHIP loci (Loh et al., 2020; Terao et al., 2020), and filtered outlier samples with multiple events (>5 sCNVs) (Fig. 1A) (Methods). sCNVs that occur early in development are clonally shared across multiple tissues and are thus expected to be present at larger cell fractions (CF) than those occurring through clonal hematopoiesis alone. Reassuringly, variants filtered as potential CHIP exhibited significantly lower CF compared to those in our final call set (Wilcoxon Rank Sum Test p = 6.4e-11) (Fig. 1B). This difference suggests that filtering reliably removes most CHIP events, though some *bona fide* sCNV may be filtered out as well, especially those coming from CN-LOH events.

**Fig. 1:**
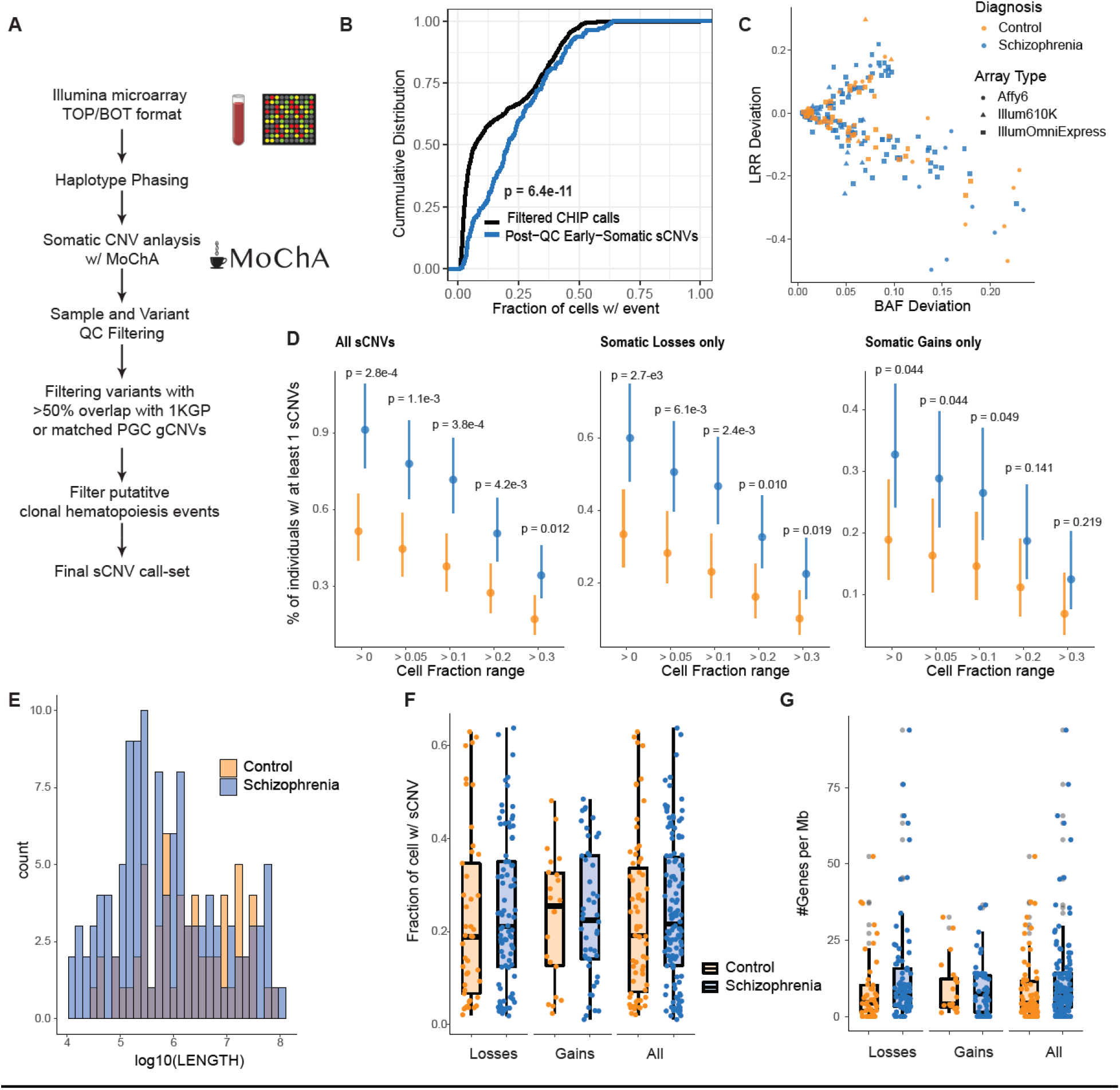
Somatic CNV burden in Schizophrenia. A) Schematic of sCNV calling and filtering strategy. B) Cumulative distributions of cell fraction with events in our final call-set (Post-QC Early developmental sCNVs), and those filtered as potential CHIP events (Filtered CHIP calls). C) Trident plot of final call-set. Each point represents an event with different colors and shapes indicating the subject’s diagnoses and array type. D) Percent of individuals with at least one sCNV in cases and controls across different minimum cell fraction thresholds. Dots represent the mean fraction, and the lines represent the 95% CI from the binomial distribution using the Wilson’s score interval with Newcombe modification. P-values were calculated using a two-sided Fisher’s Exact Test. E) Histogram of sCNV size (log10 scale) in cases and controls. F) Box plots of the sCNV cell fractions in cases vs controls. G) Box plots of the number genes overlapped per Mb of sCNVs in cases and controls.

Somatic CNVs occurred in a modest but significant fraction of SCZ cases. From the initial 13,464 SCZ cases and 12,722 controls, a total of 12,834 cases and 11,648 controls remained after QC. The final sCNV call set consisted of 197 events in 177 individuals, made up of 127 losses, and 70 gains (Table S2, Fig. 1C). These events ranged in CF from 1.10% to 63.8% (median = 21.1%), and ranged in size from 10.7 Kb to 95.3 Mb (median 686.0 Kb). The percentage of individuals with at least one sCNV was 0.91% in SCZ and 0.51% in controls (OR = 1.78; 95% Confidence Interval (CI) = 1.29-2.47; Two-sided Fisher Exact Test p= 2.68e-4) (Fig. 1C). Using the approach from Iossifov *et al*. (Iossifov et al., 2014), we obtain an ascertainment differential of 0.0091-0.0051 = 0.004. Therefore, we estimate that ∼44% (0.004/0.0091) of sCNV in SCZ contribute to the SCZ diagnosis. The sCNV incidence in controls was comparable with unaffected siblings in an earlier ASD study (0.51% vs 0.54%) (Sherman et al., 2021), while our estimates in SCZ were higher compared to ASD (0.91% vs 0.58%) (Sherman et al., 2021). This higher rate most likely reflects sensitivity improvement in the pipeline since the earlier study (Methods). To rule out potential residual CHIP events contributing to the difference in prevalence of sCNVs, we performed the burden test using different minimum cell fraction cut-offs. There remained a statistically significant enrichment in SCZ through several ranges, even when events were split into losses and gains (Fig. 1D). We further accounted for potential batch heterogeneity (Fig. S1A) using meta-analysis across each study batch containing both cases and controls, obtaining a Liptk’s combined p-value of 0.032 using a one-sided Fisher Exact Test.

In contrast with previous findings in ASD (Sherman et al., 2021), sCNVs in SCZ cases were of similar size compared to control (p = 0.26) (Fig. 1E). These events were also present at similar cell fractions in cases compared to controls (p = 0.986) (Fig. 1F). There was also no detectable difference in gene density between cases and controls (p=0.08). These trends were observed across the different batches as well (Fig. S1 B, C, D). In contrast to gCNV (Kirov et al., 2012; Marshall et al., 2017), sCNV did not show overall gene-set enrichment for the top 20% expressed brain genes (p=0.14), synaptic genes (p=0.12), or haploinsufficient genes as measured by a pLI score >0.90 (Lek et al., 2016) (p=0.54).

### Recurrent, intragenic deletions in *NRXN1* in SCZ

Some sCNV overlapped cytobands previously implicated in SCZ, but showed distinctive features. While one SCZ case had a 4.1Mb somatic deletion in cytoband 16p11.2, it was not only significantly larger than the canonical germline 16p11.2 deletions (<600 Kb) observed in SCZ and ASD (Marshall et al., 2017; Weiss et al., 2008), but also the mosaic deletion did not overlap the canonical proximal or distal events (Fig. S2A). We also observed one SCZ case with a somatic deletion in the 22q11.21 locus that was significantly smaller (686 Kb) than the recurrent germline 22q11.21 deletions observed in SCZ (2.35 Mb) (Fig.S2B). The mosaic 22q11 deletion we observed, however overlapped the genes *TBX1*, and *COMT* which have been suggested as key genes driving some of the phenotypic effects and SCZ risk of germline 22q11 deletion (Arinami, 2006; Gothelf et al., 2014).

Six individuals showed somatic deletions in cytoband 2p16.3 affecting only the *NRXN1* gene, showing remarkably stereotyped and distinctive features. The size of these events ranged from 105 Kb to 534 Kb, with CF ranging from 13.8 to 43.1%, suggesting that they occurred early in development. One deletion was limited to intron 5 (Fig. 2A), and is of uncertain disease significance since this intron also shows multiple deletions in controls in the germline (Marshall et al., 2017). In contrast, the remaining five 2p16.3 deletions had remarkably similar genic effects, removing exons 1-5 of *NRXN1*α, while leaving exon 6 and the rest of the gene intact. This stereotyped 5 exon deletion contrasts with germline deletions in *NRXN1*, previously implicated in SCZ (Flaherty et al., 2019; Marshall et al., 2017), which show highly variable breakpoints and relationships to *NRXN1* exons (Cosemans et al., 2020a; Lowther et al., 2017; Marshall et al., 2017). Therefore, the recurrent, mosaic deletion of the same exons 1-5 in all five exonic deletions would seem to demand a specific mechanistic explanation. To further assess the prevalence of somatic *NRXN1* deletions, we re-ran MoChA with a more lenient threshold and checked whether *NRXN1* CNVs identified in the original PGC study (Marshall et al., 2017) as germline might in fact be somatic. This strategy revealed a *NRXN1* deletion previously identified as germline, with an estimated CF of 41% consistent with being somatic. This variant appeared to overlap exons 4-5 for *NRXN1* (Fig. 2A), though its exact boundaries are uncertain.

**Fig. 2:**
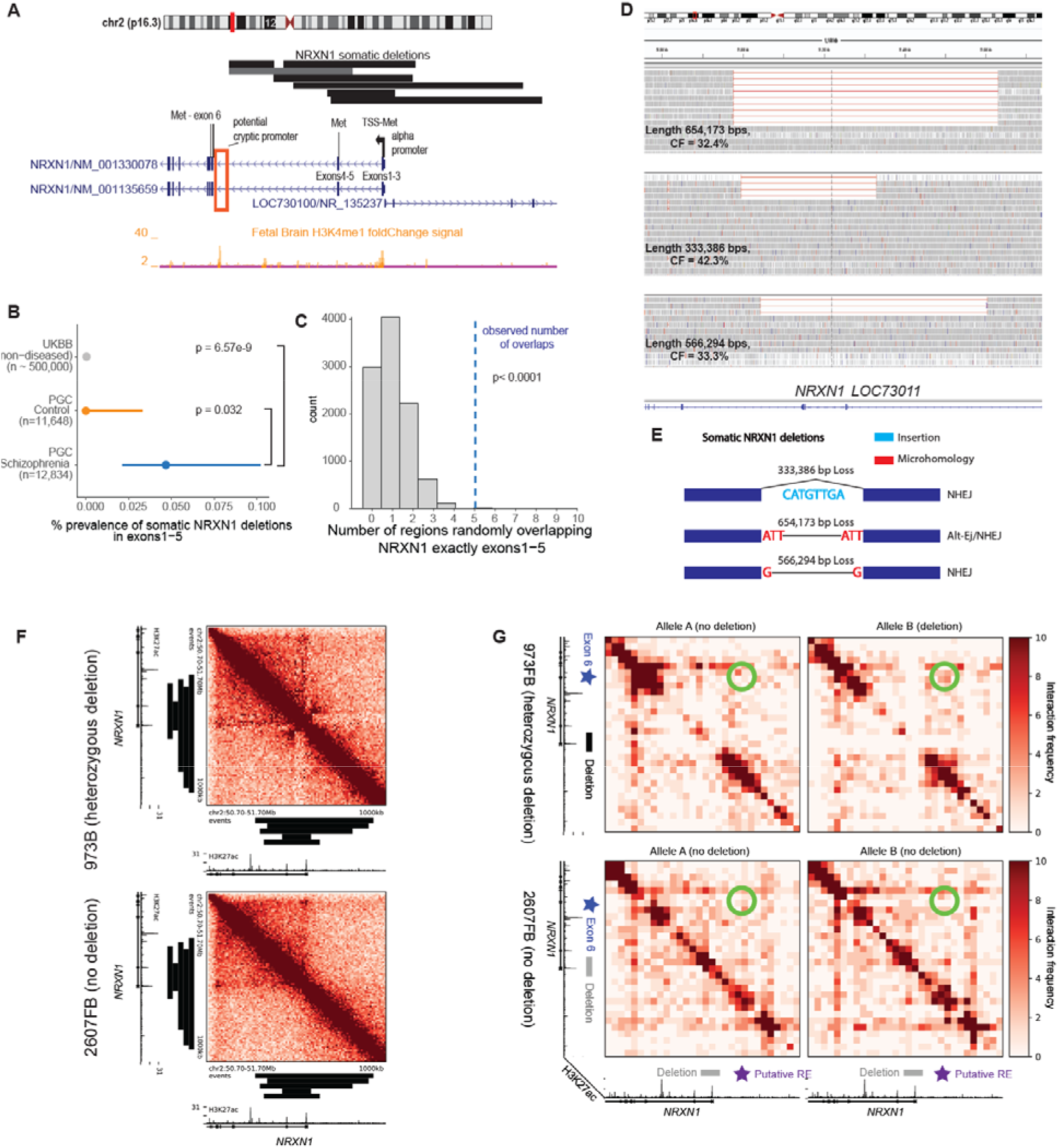
Somatic deletions of *NRXN1* exons 1-5. A) Adapted GenomeBrowser view of 7 somatic deletions of *NRXN1*. The alpha promoter and in-frame ATG/methionine sites on exons are annotated for *NRXN1*. Histone marks were obtained from Roadmap epigenomics tracks (Roadmap Epigenomics Consortium et al., 2015). Potential cryptic promoter/enhancer is marked by a red-box. Gray horizontal bar indicates CNV previously called as germline that was found to be somatic. B) Prevalence of somatic deletions of *NRXN1* exons 1-5 in SCZ, controls, and UK Biobank. P-values were estimated using two-sided Fisher’s exact test, and 95% CI were obtained using the Wilson’s score interval with Newcombe modification. C) IGV plots of the deletions of 3 SCZ subjects with somatic deletions in *NRXN1* exons 1-5 from whole-genome sequencing data. For illustration purposes not all the reads at this region were shown. D) Histogram of the distribution of number of overlaps of *NRXN1* exons 1-5 from randomly shuffling the discovered *NRXN1* sCNVs across the *NRXN1* locus. The blue dashed line is the observed number of overlaps which is equal to 6. E) Breakpoint analysis schematic showing observed insertions and microhomology at the breakpoints of the *NRXN1* sCNVs along with event length (NHEJ: non-homologous end-joining repair, Alt-EJ: alternative end joining). F) Unphased Hi-C heatmap for hiPSC derived neurons with and without 5’ (exons1-2) deletions. Black bars indicate regions of somatic *NRXN1* deletions. G) Phased Hi-C heatmaps for hiPSC derived neurons. Green circles indicate areas of higher signal with 5’ deletion of *NRXN1* in the affected allele. Black bar indicates germline *NRXN1* deletion of exons1-2. RE stands for regulatory element.

Comparing the burden of *NRXN1* somatic deletions in our SCZ cases vs controls revealed a significant enrichment in cases (Two-sided Fisher Exact Test p = 0.032 (exonic only), p = 0.016 (exonic + intronic); Fig.2B). Using previously generated sCNV calls from the UK Biobank (Loh et al., 2018, 2020), we identified two persons without history of psychiatric disorder out of ∼500,000 individuals with similar sCNV breakpoints affecting exons 1-5 in *NRXN1*. Although the arrays used in the UK Biobank have different sensitivity compared to the arrays used in this study, they should have comparable sensitivity to detect these large events at CF >10% (Loh et al., 2018). Consequently, while we cannot fully rule out batch effect bias, combining our results with the UKBB suggest an enrichment of exons1-5 *NRXN1* deletions in the somatic state in SCZ (OR=117.08; 95% CI = 20.91-1165.84; Fisher Exact Test p=6.57e-9; Fig. 2B). To further assess whether we could have observed 5 overlaps on exons1-5 by chance, we randomly shuffled the 7 *NRXN1* sCNV regions we discovered across the *NRXN1* locus and computed the number of overlaps, showing that observing 5 overlaps of exactly exons1-5 was an extremely unlikely event (p<0.0001, Fig. 2C). Remarkably, a similar study with a similar pipeline and dataset as this study (Sherman et al., 2021) on ASD and control samples did not detect somatic deletions in *NRXN1* overlapping exons1-5, suggesting specificity of this event to SCZ.

We were able to obtain 40X whole genome sequencing (WGS) from 3 cases with *NRXN1*α deletions processed at the Broad Institute, confirming that each event removed exons1-5 of the gene with estimated CFs of 42.4%, 33.3%, and 32.4%, as expected (Fig. 2D), and defining their breakpoints at basepair resolution. WGS analysis showed that none of the *NRXN1* sCNVs breakpoints were recurrent, nor overlapped known interspersed repeats or low complexity DNA sequences.

Further breakpoint analysis of these *NRXN1* sCNVs using previously established classification criteria (Kidd et al., 2010; Yang et al., 2013) (Fig. 2E) suggested diverse mechanisms of formation. One event had only 1 bp of microhomology (MH) suggesting that this event arose via non-homologous end joining repair (NHEJ). Another event had a 3 bp MH implicating an alternative end-joining repair mechanism (alt-EJ). The last event had no MH but revealed a 8bp insertion at the breakpoint. This insertion is small enough to have occurred due to non-template directed repair associated with NHEJ, although it is also possible that a fork-stalling template switching mechanism might have occurred as well, but this mechanism tends to produce insertions > 10bp and usually occurs where some microhomology exists at the ends (Yang et al., 2013). Taken together these results suggest that the somatic deletions of *NRXN1* that we observed do not show recurrent breakpoints due to instability of the genomic region around exons 1-5.

### *NRXN1* deletions suggest a potential cryptic promoter in human induced neurons

The absence of a genomic mechanism for the recurrent somatic deletions in *NRXN1*α suggests the alternative hypothesis that the recurrence reflects some unknown but specific effect of these deletions on *NRXN1* gene function. These sCNVs overlap the *NRXN1*α promoter along with the first in-frame ATG transcription start site, which would be expected to disrupt transcription of the full alpha isoform from that allele (Fig. 2A), while leaving downstream beta and gamma isoforms intact, since they initiate transcription further downstream. Intriguingly, the somatic deletions leave intact H3K4Me1 histone marks that lie just 5’ from exon 6, which contains an in-frame ATG (Fig. 2A). These features might be indicative of the presence of a cryptic promoter or enhancer adjacent to the in-frame ATG in exon 6, potentially producing a N-terminal truncated *NRXN1*α for deletions overlapping exons 1-5. This truncated protein would lack the signal peptide required for shuttling to the cell surface, potentially causing abnormal trafficking. Similar germline *NRXN1* deletions have been shown to cause accumulation of the *NRXN1* intracellular binding protein CASK in human induced pluripotent cells (iPSC) from SCZ patients (Pak et al., 2021).

To further explore the potential functional role of somatic deletions in the 5’ end of *NRXN1*, we generated Hi-C data from neurons differentiated from human iPSC containing heterozygous germline deletions in the 5’-end (exons 1-2) and compared them to an iPSC line that had no germline deletion in *NRXN1* (Methods). Unphased Hi-C heatmaps in iPSC-neuron showed that somatic deletions affecting exons 1-5 all fully overlap the topologically associating domain (TAD) co-localized with the alpha promoter (Fig. 2F). Recently, disruption of TAD boundaries by germline structural variants have been associated with developmental disorders, as well as SCZ (Bompadre and Andrey, 2019; Halvorsen et al., 2020). These observations together suggest that 5’ *NRXN1* deletions might disrupt the structural integrity of the TAD boundary in SCZ and could result in ectopic enhancer-promoter miswiring and dysregulated gene expression.

To investigate possible 3D genome miswiring due to *NRXN1* deletions, we generated allele-specific, phased Hi-C maps in both control as well as deletion-carrying SCZ iPSC-neurons (Methods). Surprisingly, we observed the *de novo* formation of an ectopic looping interaction (Fig. 2G, green circle) between exon6 of *NRXN1* (Fig 2G, blue star) and a putative non-coding cis regulatory element upstream of the *NRXN1* alpha promoter (Fig 2G, purple star). This ectopic loop appeared to be specific to the deletion-harboring allele of the sample bearing a heterozygous deletion spanning the alpha promoter at the 5’ end of *NRXN1* (973FB) and was not observed on either allele in samples that lacked the deletion (2607FB). Because the interaction spans the deleted region, we hypothesize that the deleted region contains an element with some degree of boundary function which prevents this loop from forming under normal circumstances. Consistent with our hypothesis, the frequency of non-specific interactions increased across the boundary only on the *NRXN1* deleted allele, which is indicative of allele-specific severe compromise of TAD structural integrity in SCZ (Fig. 2G). Together, our working model is that the *de novo* looping interaction in 5’ *NRXN1* deletions in SCZ connecting exon6 to a putatively regulatory element could promote spurious pathological transcripts initiating at exon 6.

### Recurrent sCNVs in the *ABCB11* gene in treatment-resistant SCZ cases

We identified 6 SCZ cases with focal sCNVs in the *ABCB11* gene (five deletions and one gain; Fig. 3A), which has been previously associated with anti-psychotic response (Gonzalez-Covarrubias et al., 2016; Vita et al., 2019). These events were all smaller than the average sCNVs we observed, with sizes ranging from 10.5 Kb to 35.4Kb, and with CFs ranging from 18.3% to 26.8 %, suggesting that these events occurred early in development. *ABCB11* encodes a member of the superfamily of ATP-binding cassette (ABC) transporters, which has a key role of transporting proteins across the cell-membrane using a “hinge” mechanism (Gonzalez-Covarrubias et al., 2016) in hepatocytes, the cells involved in a wide range anti-psychotic metabolism. All the sCNVs in the *ABCB11* gene overlapped the ABC transporter 1 domain and the domain responsible for interaction with the HAX1 protein (Fig. 3B). HAX1 aids in the internalization of the ABCB11 transporter through clathrin mediated endocytosis (Alogaili et al., 2020; Ortiz et al., 2004). Consequently, it is expected that deletions might not only alter the protein’s function by altering the transporter domains, but also prevent the removal of ABCB11 from the cell surface, potentially leading to a dominant negative loss of function. Since the sCNVs in *ABCB11* do not overlap the gene’s promoter, and there are in-frame ATG sites in downstream exons 19 and 20, a truncated protein could be produced. The consequences of the somatic duplication event are less clear. We also note that 4 out of the 5 deletions and the duplication event overlap one of the transmembrane domains, further supporting the idea that these sCNVs might have a detrimental effect on ABCB11 function. There was a significant enrichment of *ABCB11* sCNVs in cases compared to controls (Two-sided Fisher Exact Test, p = 0.03 Fig. 3C).

**Fig. 3:**
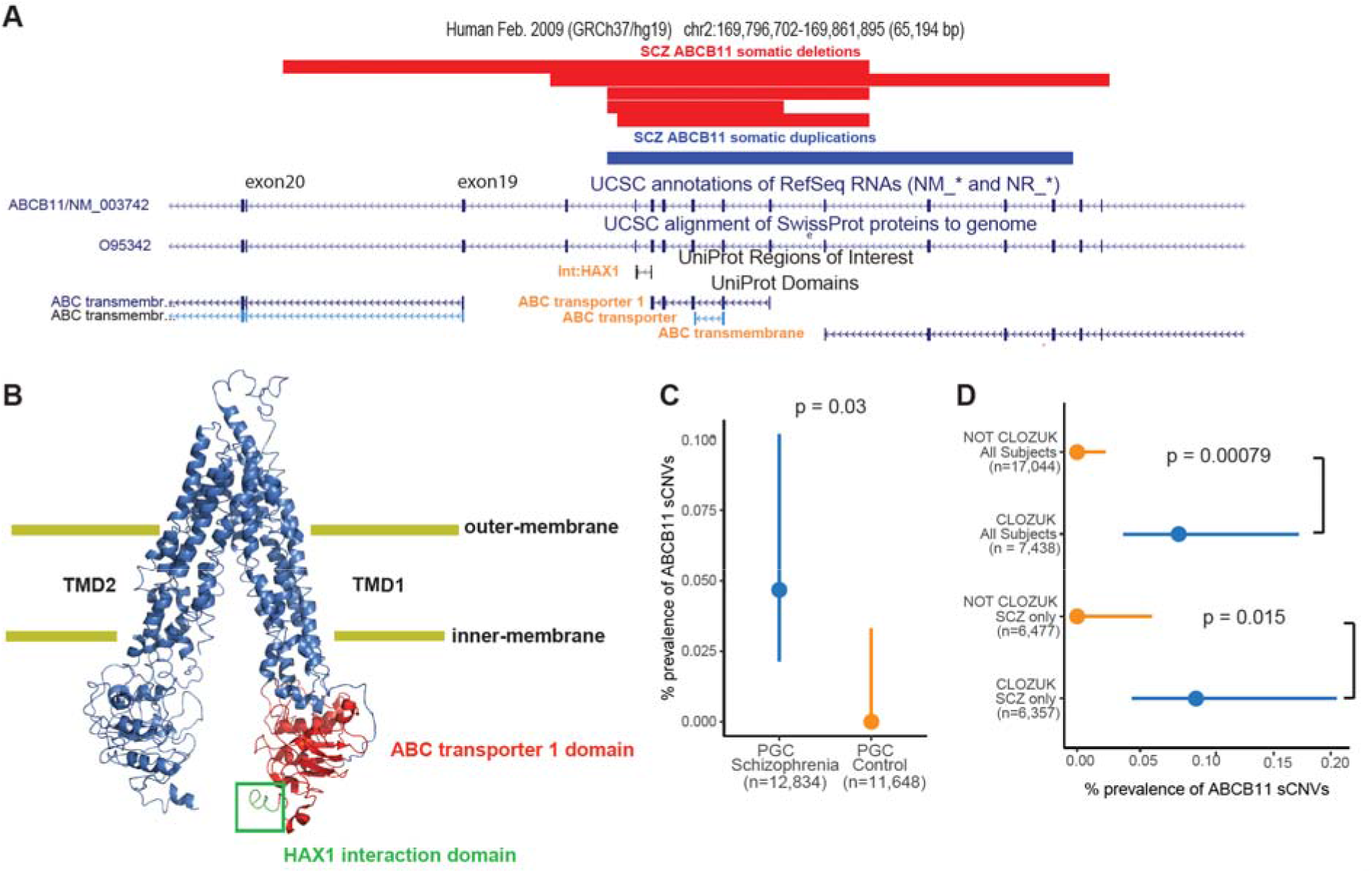
Somatic CNVs overlap *ABCB11* gene in treatment-resistant SCZ subjects. A) Adapted GenomeBrowser view of 5 somatic deletions and 1 somatic duplication of *ABCB11*. Protein domains of interest overlapped by the sCNVs have orange font. B) PyMOL schematic of the ABCB11 protein indicating the HAX1 protein interaction region and the ABC transporter 1 domain which are affected by the somatic deletions of *ABCB11*. The protein is on a “inner-open” conformation since it is not bound to ATP. C) Prevalence of intragenic sCNV in *ABCB11* gene in SCZ and controls. D) Prevalence of intragenic sCNV in *ABCB11* gene in CLOZUK cohort samples. For C and D, p-values were estimated using two-sided Fisher’s exact test, and 95% CI were obtained using the Wilson’s score interval with Newcombe modification.

Further inspection revealed that all 6 cases with *ABCB11* sCNV came from batches of CLOZUK (Consortium et al., 2014), a treatment-resistant schizophrenia (TRS) cohort. The samples from these patients were obtained from individuals that had received a diagnosis of TRS and were taking clozapine and thus were subject to standard blood monitoring for this drug (Hamshere et al., 2013). Even though the CLOZUK samples constituted a significant portion of our study, observing 6 cases from only this cohort represents a statistically significant enrichment (Two-sided Fisher Exact test: p = 0.00079, and p = 0.015 for and SCZ only, respectively; Fig. 3D). *ABCB11* sCNVs were not found in any previous analyses of healthy individuals from the UK Biobank and Biobank Japan (Loh et al., 2020; Terao et al., 2020). The high cell fraction of these events suggests an early-developmental origin of these mutations which might have predisposed these SCZ patients to develop treatment resistance to anti-psychotics. Thus, while these variants might not have been directly implicated in SCZ liability, they might have influenced the patients’ clinical management. Out of the samples that had *ABCB11* sCNV, only 2 (1 gain and 1 loss) were available for WGS. Unfortunately, the predicted breakpoints fall in repetitive regions of the genome (SINE elements) (Fig. S3), making it difficult to identify exact breakpoints, though the presence of these repetitive sequences suggest a potential mechanism of somatic deletion through microhomology.

Combining the *ABCB11* somatic deletions we observed in our SCZ cases with germline deletions identified as part of the phase 2 PGC germline CNV dataset revealed robust overlap between the mosaic deletions we detected and those present in separate SCZ cases in the germline state. There were 5 SCZ cases with gCNVs at the *ABCB11* locus, with three of them coming from the CLOZUK cohort (Fig. S4). We were not able to obtain clinical data whether the remaining two cases had TRS. Interestingly, there were 6 control subjects with germline deletions in *ABCB11*, but these events tended to cluster downstream from the SCZ gCNV and sCNV variants (Fig. S4). SCZ risk association analyses combining germline and somatic deletions of *ABCB11* revealed statistically significant association at the HAX1 interaction site and ABC transporter 1 site (peak association p = 1.4e-4). While not genome-wide significant (p = 8.3e-8), it suggests a potential role of *ABCB11* in treatment response in SCZ.

### Potential of sCNVs to implicate novel genomic regions in SCZ

Comparison of the genomic features of gCNVs and sCNVs suggest that sCNVs contribute to risk by distinct molecular mechanisms. We obtained previously identified rare (minor population allele frequency <0.5%) gCNV calls of SCZ cases from the arrays used in our current study (Marshall et al., 2017). Compared to these rare gCNVs, sCNVs were larger (Fold-Change = 4.57, 95% CI = 3.76-5.48, mixed-effect log-normal regression p < 2e-16) and involved more genes (Fold-Change = 1.84, 95% CI = 1.51-2.23; mixed-effect log-normal regression p=4.45e-9) (Fig. 4A-B). We observed that genomic regions affected by rare gCNVs present in at least 5 SCZ cases overlapped 43.6% of all the gCNVs, whereas these regions overlapped only 4.48% of SCZ sCNVs (Fig. 4C). This difference in genomic regions persisted throughout for rare gCNVs present at different minimum recurrence cut-offs (Fig. 4C). These findings suggest that, with sufficient statistical power, mosaic events might offer new insights into different risk regions of the genome as well as mechanism of disease.

**Fig. 4:**
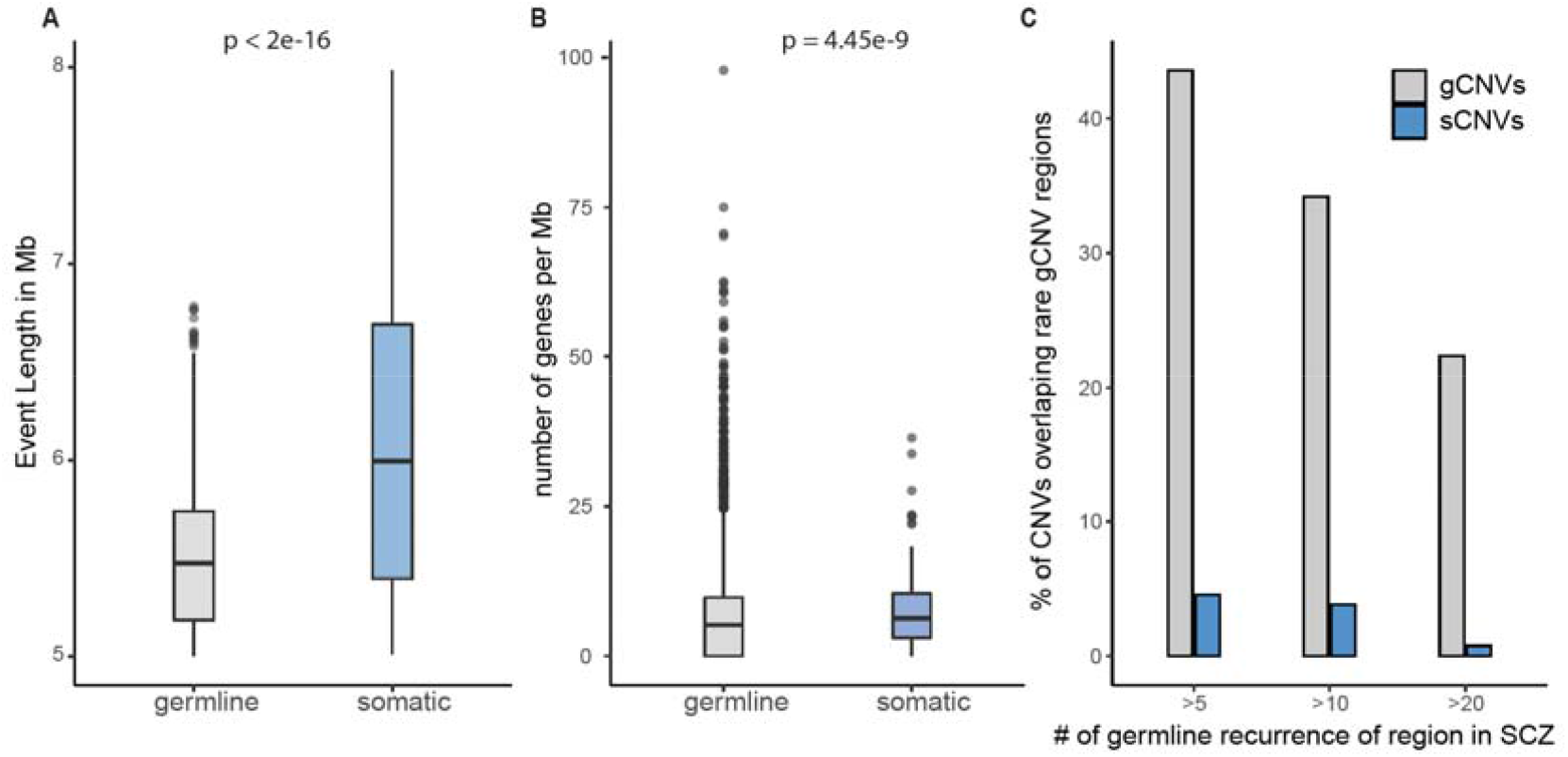
Somatic CNVs differ in size, gene content, and location from germline CNVs in SCZ. A) Boxplot of event length in SCZ in the somatic and germline state. B) Plot of the number of genes affected per Mb. The p-values for panels A and B were calculated using mixed effect model regression with batch as a random effect. C) Bar plots showing the percentage of the CNVs on each category that overlapped recurrent germline rare CNV regions in SCZ, across three different minimum recurrence thresholds.

## Discussion

We show that somatic CNVs contribute a modest but significant part of the genetic architecture of SCZ, mirroring previous findings on rare germline and *de novo* CNVs (Kirov et al., 2012; Marshall et al., 2017). The sCNV excess burden of 0.4% in SCZ likely represents a lower bound, since we are limited to detecting events with large enough cell fractions to be present as mosaics in different tissues such a blood, and are not able to assess those events that might be private to the brain.

In this study we also report the discovery of 5 SCZ cases with mosaic deletions of exons 1-5 that also cover the promoter of *NRXN1*α. Deletions of these exons were present in only 2 out ∼500,000 individuals in the UK Biobank, which has an ascertainment bias for healthy individuals, and were absent from our control cohort. This high prevalence in our SCZ cohort for relatively large ∼100Kb-500Kb events suggests that mosaic deletions of exons 1-5 might contribute to SCZ risk.

A study characterizing germline *NRXN1* deletions from 19,263 clinical arrays in individuals with neurodevelopmental disease found that most of these events were present in the 5’ end of *NRXN1* and covered exons 1-5 (Lowther et al., 2017). Our group published a case series that suggested that deletions in *NRXN1* predispose individuals to severe developmental disorders through inherited CNVs (Ching et al., 2010). In that study, two subjects with severe developmental delay had inherited deletions of exons 1-5. In contrast, germline deletions of *NRXN1* in SCZ are widely distributed throughout the gene (Marshall et al., 2017), rather than being concentrated in the first few exons as in neurodevelopmental disorders (Cosemans et al., 2020b; Lowther et al., 2017). This contrast might indicate that germline deletions of exons 1-5 might result in more severe developmental phenotypes, but if present in only a fraction of cells, it would result in a milder phenotype resembling SCZ.

These developmental and neuronal abnormalities can be partially explained by the effect that 5’ deletions involving exons 1-5 and the *NRXN1*α promoter might have on *NRXN1* function. A recent study characterized the neuronal impact of aberrant *NRXN1*α splicing using hiPSC derived neurons (Flaherty et al., 2019). The 5’ deletions were associated with decrease in the *NRXN1*α isoform and an increase of *NRXN1*β. Heterozygous hiPSC neurons had a reduction in mature neurons and decreased spikes compared to controls, as well as decreased neurite number and total length. Taken together, these data suggest that deletions of exons 1-5 of *NRXN1* can lead to severe developmental abnormalities in the germline state by disrupting neuronal maturation and function.

While the most parsimonious model of pathogenicity of somatic deletions in *NRXN1* exons1-5 is simple loss-of-function through deletion of the alpha promoter, the vast diversity of *NRXN1* isoforms warrants further exploration of alternative mechanisms. Our analysis of Hi-C data using the same hiPSC neurons from Flaherty *et al*., suggests a potential formation of a cryptic promoter once the *NRXN1* alpha-promoter is deleted, potentially forming an N-terminal truncated form of *NRXN1*, leading to a novel dominant negative mechanism by trapping *NRXN1*α in the cytoplasm. This mechanism is consistent with higher intracellular protein levels of a NRXN1-binding protein CASK in human iPSC lines from SCZ patients with 5’ *NRXN1* deletions (Pak et al., 2021). This potential cryptic promoter might have been missed in previous studies due to the difficulty of developing targeted transcript primers not anchored at the 5’ end (Flaherty et al., 2019). Further transcriptional and functional experiments could better validate the presence and role of this putative cryptic promoter in *NRXN1* and SCZ biology.

In this study we also found 5 early developmental recurrent somatic deletions in the *ABCB11* transporter gene. These deletions were present only in the SCZ cases diagnosed with treatment-resistant schizophrenia, which is defined as nonresponse to at least two antipsychotic medications (National Institute of Health and Clinical Excellence, 2014), and affects ∼30% of individuals with SCZ (Meltzer, 1997). Genes in this transporter family, including *ABCB11* have been previously associated with differential response to anti-psychotics (Vita et al., 2019). However, the exact mechanism by which mutations in these genes might lead to poor response to anti-psychotics remains unknown.

The recurrent somatic deletions in *ABCB11* suggest a dominant negative genetic mechanism. The *ABCB11* gene is not dosage sensitive as measured by having a low pLI score ∼0 (Lek et al., 2016), suggesting that a dominant negative mechanism is required for heterozygous mutations to have an effect of phenotype. The somatic losses of *ABCB11* deleted the region that encodes for the protein domain responsible for interaction with HAX1. Disruption of this interaction prevents the ABCB11 transporter from being recycled, leading to an increase of ABCB11 on the cell surface (Alogaili et al., 2020; Ortiz et al., 2004). Since the somatic deletions also affect parts of the transmembrane and transporter domain of the protein, it follows that these heterozygous sCNVs might cause a dominant negative phenotype by persistently expressing altered ABCB11 transporter proteins in the cell’s apical surface. Taken together these data suggests that early developmental somatic losses in *ABCB11* might predispose a subset of SCZ patients to have poor response to anti-psychotics. Future functional studies are needed to validate this potential dominant negative mechanism, and test the effect of disrupting HAX1 interaction in anti-psychotic response.

In summary, somatic CNVs in SCZ tend to be more prevalent compared to controls, suggesting a potential for sCNVs to contribute to disease liability and affect treatment response. These data suggest a modest but potentially important role of sCNVs to the genetic architecture of SCZ.

## Data Availability

All data produced in the present work are contained in the manuscript. Individual level SNP-array data is part of the Psychiatric Genomic Consortium with the corresponding privacy agreement. Access can be provided by applying through this website (https://www.med.unc.edu/pgc/shared-methods/how-to/)

## Acknowledgements

E.A.M. is supported by the Harvard/MIT MD-PhD program (T32GM007753), the Biomedical Informatics and Data Science Training Program (T15LM007092), and the Ruth L. Kirschstein NRSA F31 Fellowship (F31MH124292). G.G. is supported by NIH grant (R01HG006855), NIH grant (R01MH104964), and the Stanley Center for Psychiatric Research. S.A., A.C, and C.A.W were supported by the NIMH grant (U01MH106883) through the Brain Somatic Mosaicism Network (BSMN). C.A.W is an Investigator of the Howard Hughes Medical Institute. C.A.W and E.A.L are supported by the Allen Frontiers Program through the Allen Discovery Center for Human Brain Evolution. E.A.L. is supported by the NIH grants (K01 AG051791, DP2 AG072437, R01AG070921) and SUHF foundation. J.S. is supported by NIH grants (MH113715, MH119746, MH109501, MH119746). J.E.P-C and K.J.B. are supported by a Chan Zuckerberg Institute grant (2020-221479). J.E.P-C is supported by NIH grants (DP1OD031253, R01NS-114226, R01MH12026,U01DK127405).

## Contributions

E.A.M. and C.A.W. conceived and designed the study. E.A.M. designed and implemented the statistical methods. E.A.M performed computational analyses, with assistance from M.A.S. and G.G.. J.S. curated the data and facilitated access. S.M. and A.C. facilitated acquisition of samples for whole-genome sequencing validation. J.T.R.W., M.O., and P.S. facilitated clinical and genomic data procurement for validation and interpretation. P.R., S.A., and K.J.B. generated the Hi-C data. T.G.G., J.E.P-C analyzed and interpreted the Hi-C data. E.F. and K.J.B. generated and characterized the human iPCs/neurons. E.A.L., P.-R.L., S.A.M., and J.S. provided comments and guidance throughout. E.A.M., E.A.L, and C.A.W. wrote the manuscript.

## METHODS

### SNP array data acquisition

Allelic intensity data for cases and controls were obtained from the Psychiatric Genomic Consortium (PGC) CNV working group. The exact details of the data generation were previously described (Marshall et al., 2017). SNP array data consisting of 13,464 SCZ cases and 12,722 controls was obtained. These data were profiled with the Illumina OmniExpress, OmniExpress plus exome chip, Illum610K, and Affymetrix SNP6.0 arrays. For each determined position the B allele frequency (BAF; proportion of B allele), Log-R ratio (LRR; total genotyping intensity of A and B alleles), and genotype calls, were calculated.

### Data processing

The genotypes from the SNPs from the arrays were phased using the Eagle2(Loh et al., 2016) software. Then, the BCFtools plug-in MoChA (2021-01-20 release) was used to confidently call mosaic CNVs, by taking advantage of long-range haplotype phasing of heterozygous SNP sites and BAF estimates of genotype array data. Genotyping and intensity data from Illumina platforms were distributed by the PGC in the Illumina GenomeStudio Final Report format, with the genomic positions genotyped using the hg18 human reference genome. To convert the Final Report format to VCF format, the rsID numbers were used to liftover coordinates to hg19, discarding positions without rsID, similar to Sherman et al(Sherman et al., 2021). Costum scripts were used to transform Final Reports to BCF format, and Illumina’s TOP-BOT format was converted to dbSNP REF-ALT format using a modified version of BCFtools plug-in fix-ref. MoChA calculates cell fraction from BAF as follows:

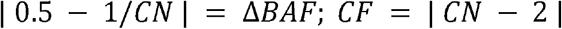

where CN is the copy number and ΔBAF is the deviation of B allele fraction compared to 0.50. This equation is valid for gains and losses.

### Variant Level Quality Control

In accordance with the suggestions of the MoChA processing pipeline, the following variants were filtered out: more than 2% genotypes missing, evidence of excess heterozygosity (p < 1e-6, Hardy-Weinberg equilibrium test), correlation of autosomal genotypes with sex (Fisher exact test comparing number of 0/0 genotypes vs number of 1/1 genotypes in males and females), variants falling within segmental duplications with low divergence (< 2%). This variant-level QC was performed on each separate batch.

### Sample-Level Quality Control

In order to filter out samples with contamination from another individual two statistics were calculated: BAF concordance and BAF autocorrelation. Briefly, BAF concordance calculates the probability that an adjacent heterozygous SNP has a deviation from a BAF of 0.5 given that the previous heterozygous site had the same deviation from 0.5(Vattathil and Scheet, 2013). BAF autocorrelation is the correlation of the BAF statistic at consecutive heterozygous sites once adjusted for the genotype phase. Samples with contamination with DNA from another individual would be expected to have a BAF concordance > 0.5 and BAF autocorrelation > 0 because of allelic intensities correlated at variants within haplotypes shared between sample DNA and contaminated DNA. Samples with BAF concordance > 0.51 or BAF autocorrelation > 0.03 were removed.

### Event type classification

An Expectation-Maximization algorithm was applied to classify events as either a Gain, Loss, or CN-LOH. The algorithm determines the slopes that characterizes the relationship between the deviation of the LRR from 0 |ΔLRR|, and the BAF deviation from 0.5, |ΔBAF|. In other words, the events are classified based on the optimization of linear regression parameters described by |ΔLRR| = |Δ*BAF*|β_C_ + ∈, where *c* ∈ {*Gain,Loss,CN - LOH*], *β*_*C*_ is the slope for each event type, 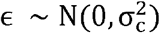 is the error for each event-type clustering.

To further enhance the robustness of the classification method, we used the fact that CN-LOH events are expected to be less common within the chromosomes compared to events that extend to the telomeres. Since CN-LOH events are thought to arise during mitotic recombination, for them to occur within a chromosome would require a double crossover, which is highly unlikely. To incorporate this information into the classification model, we estimated the frequency using the UK Biobank sCNV calls(Loh et al., 2018, 2020) for of each event type occurring on telomeres and interstitially. These frequencies were used as priors to multiply the likelihoods for each event type, resulting in posterior probabilities. The computation for each event *S*_*i*_ is as follows: Let *X*= |Δ*BAF*| and *Y*= |Δ*LRR*|, then 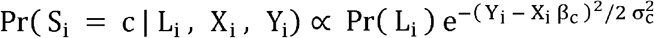, where *L*_*i*_ is an indicator of whether the event involves a telomere, and c is defined as above. This estimation is calculated for each event type and then normalized to sum to one.

### Filtration of Mosaic CNV calls

Filtration was focused on removing potential germline events and events likely to arise due to age-related clonal hematopoiesis, as well as artifacts. We required events to have a log10-odds >10 for the model based on BAF and phase, which measures how much more likely the data for a given segment of DNA is consistent with a non-diploid model than a diploid model. Events that were classified as copy number polymorphism (known CNV polymorphisms in 1000 Genomes Project) by MoChA were filtered out as possible germline events. We further excluded events that had a reciprocal overlap with events from control samples or with any CNVs reported in the 1000 Genomes project by >50%. Events that overlapped >50% with germline events previously identified in the same sample by the PGC (Marshall et al., 2017) were also removed for duplications, since small duplications with high BAF deviations can be mistakenly identified as somatic variants. Copy number state was taken into consideration when calculating overlaps, i.e. overlap between gains and losses were not considered. Calls with an estimated cell fraction of 1 were also removed. For gains, we further removed any events with a deviation in BAF greater than 0.10 to have a conservative assurance that germline gains were not misclassified as mosaic, as germline gains tend to be small and produce large deviations from the a BAF of about 1/6 (Loh et al., 2018).

Finally, since moust our datasets did not include age information for individuals besides the broad estimate of being younger than 40, we used a conservative approach to remove events that could have risen from clonal hematopoiesis. CN-LOH events were fully excluded from any downstream analysis as these events have been shown to be largely enriched in clonal hematopoiesis events(Loh et al., 2018). We also removed sCNVs that contained loci commonly altered within the immune system, specifically IGH (chr14:105,000,000-108,000,000) and IGL (chr22:22,000,000-40,000,000). We also excluded CNVs within the extended MHC region (chr6:19,000,000-40,000,000). In addition, we removed deletion involving the following loci that are frequently affected by clonal hematopoiesis: 20q11, *DNMT3A, TET2*, 13q14, 17p, 5q14, *ATM*. We removed duplications in 15q. We also removed any sCNVs in 7q34 and 14q11.2, as well as trisomy 12 events. We also removed events whose copy-number state could not be determined.

### Overall Burden Analysis

To test the hypothesis of whether more individuals with at least one sCNV of cell fraction greater than a given cell fraction cut-off in cases vs controls, the two-sided Fisher’s Exact test was used (Sherman et al., 2021). The 95% confidence intervals were calculated using Wilson’s score interval. For the meta-analysis using each batch separately we used a one-sided Fisher’s Exact test. The p-values were combined using the Tippet’s (minimum p-value), and the Liptk’s (weighted sum of p-values) approaches.

### Cell fraction, geneset, length, and gene number burden analysis

To calculate the contribution of the features of gene, length, and gene number burden, we fit a mixed effect logistic regression on the case-control phenotype as the outcome variable. Let *y*_*i*_ ∈ {0,1] be an indicator of whether the subject is diagnosed with SCZ or a control respectively. We modeled the burden as follows:

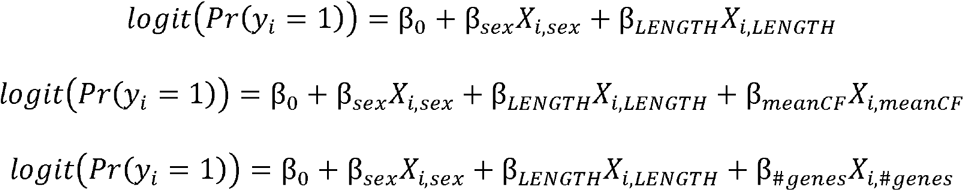

where *X*_*LENGTH*_ and *X*_*#genes*_ are the sum of the length and number of genes overlapped by events of individual i, and *X*_*meanCF*_ is the mean cell fraction of the events of individual i. Inference was not altered by the sufficient statistic used to summarize cell fraction (i.e. min, max, median). In the models above we were interested on testing whether β≠ 0 for the feature of interest. The models were fit using a generalized mixed-effect model as implemented by the R package lme4 (Bates et al., 2015) to account for the sample collection batches of the PGC. Statistical significance was assessed using the Satterwhite approximation to the t-test as implemented in the package lmerTest (Kuznetsova et al., 2017).

### Gene-set Enrichment analysis

We used a similar approach as recommended by Raychoudhri et al (Raychaudhuri et al., 2010) to control for event length and rate, which might result in false positive associations with neuronal genes. Namely, we fit the following model

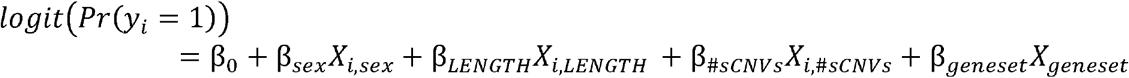

where the parameters are as defined the section above, but with *X*_*#sCNVs*_is the number of sCNVs in that individual, and *X*_*geneset*_ is the number of genes in an event that intersect a gene-set of interest. We then used the likelihood ratio test to test whether *β*_*geneset*_ ≠ 0. We used 3 gene-sets: (1) Brain expressed genes: defined as the top 20% of brain expressed genes from the GTEx GTEx_Analysis_2017-06-05_v8_RNASeQCv1.1.9_gene_median_tpm.gct.gz (https://www.gtexportal.org/home/datasets). (2) Synaptic genes obtained from SynaptomeDB (http://metamoodics.org/SynaptomeDB/index.php). (3) High pLI genes, i.e. pLI > 0.90, obtained from ExAC (file: fordist_cleaned_nonpsych_z_pli_rec_null_data.txt) (https://gnomad.broadinstitute.org/downloads).

### Permutation test for enrichment of sCNV overlapping exons 1-5 of NRXN1

We used the R package regioneR (Gel et al., 2016) to randomly shuffle the 7 sCNV that overlapped *NRXN1* across the *NRXN1* locus using the randomizeRegions function. We added a padding of 1Mb to the 5’ and 3’ ends of the *NRXN1* locus. After randomly shuffling the sCNV we counted how many segments overlapped exons 1-5. We repeated this procedure 10,000 times. To calculate a p-value we obtained the fraction of overlaps greater than the observed 5. Since we performed 10,000 iterations our smaller possible p-value was 0.0001.

### Breakpoint microhomology analysis

For the *NRXN1* somatic deletions, we identified the breakpoints at the single base resolution by looking for clipped reads with IGV (Thorvaldsdóttir et al., 2013) in the vicinity of discordant paired reads mapping to genomic locations that implied a larger insert size than expected. Microhomology was identified by looking at the surrounding bases of the clipped reads covering the breakpoint and looking for corresponding identical basepairs.

Characterization of the mechanism of origin was identified using the strategy described in Yang *et al* (Yang et al., 2013). In brief, if there was no microhomology nor insertions >10 bp, the event was predicted to be created by non-homologous end joining repair (NHEJ). If there was a microhomology >2 bp but <100 bp, the event was classified as alternative end joining (alt-EJ). If the microhomology was >100bp, which was not observed in this study, the event was classified as non-allelic homologous repair (NHAR).

The cell fraction of the events was estimated by identifying the breakpoints as above, and counting the number of clipped reads supporting the breakpoints from IGV images. Specifically, the number of clipped reads was divided by the sequencing depth at that site and multiplied by 2. For each event, the estimate of the cell fraction was obtained from the breakpoint with the highest coverage.

### Germline CNV Analyses

We obtained gCNV final calls from the SCZ Phase 2 study by the PGC CNV working group(Marshall et al., 2017). We narrowed down the gCNV calls to those that were identified in the same genotype arrays that were analyzed for sCNVs. To further control for sensitivity between the methods used to call sCNVs and gCNVs we focused on gCNV events with size >100Kb. Length and genic burden analyses were performed using a mixed effect model framework using sample batch as the random effect.

### *In situ* Hi-C from hiPSC-derived neurons

Forebrain neurons were generated as previously described (Flaherty et al., 2019). Briefly, neural precursor cells (NPCs) derived from hiPSCs with heterozygous germline deletions in the 5’-end (exons 1-2), 3’-end (exons 21-23) and from an hiPSC line with no germline deletion in NRXN1 were seeded at low density and cultured in neural differentiation medium (DMEM/F12, 1xN2, 1xB27-RA, 20 ng ml−1 BDNF (Peprotech), 20 ng ml−1 GDNF (Peprotech), 1mM dibutyryl-cyclic AMP (Sigma), 200nM ascorbic acid (Sigma) and 1 μgml−1 laminin (ThermoFisher Scientific) 1–2 days later. Cells were maintained in differentiation medium for 7.5 weeks before harvesting.

*In situ* Hi-C libraries were generated from 500K-1 million cultured hiPSC-derived neurons using the Arima Hi-C kit (Arima Genomics, San Diego) per manufacturer’s instructions without modifications. Briefly, in situ Hi-C consists of 7 steps: (1) crosslinking cells with formaldehyde, (1) digestion of the DNA using a proprietary restriction enzyme cocktail within intact nuclei, (3) filling and biotinylation of the resulting 5’-overhangs, (4) ligation of blunt ends, (5) shearing of the DNA, (6) pull down of the biotinylated ligation junctions with streptavidin beads, and (7) analyzing these fragments using paired end sequencing. The resulting Hi-C libraries were sequenced on the Illumina HiSeq1000 platform (125bp paired-end) (New York Genome Center).

### Hi-C read alignment

Hi-C reads were aligned to the hg19 reference genome using bwa mem (v0.7.17-r1188) using the flags “-SP5M” (“-SP” for aligning each end of the paired end reads separately, “-5” to force always reporting the 5’ part of a chimeric read as primary).

Aligned reads were subsequently used for two different tasks: 1) variant calling with the GATK pipeline followed by HapCUT2 phasing, and 2) Hi-C matrix construction via pairtools.

### Preprocessing for variant calling

Duplicate Hi-C reads were marked using Picard’s MarkDuplicates (via GATK, v4.0.12.0). Bamfiles were recalibrated using the GATK BQSR (base quality score recalibration) procedure. Briefly, BaseRecalibrator was run using dbSNP build 138, the Mills + 1000 Genomes gold standard indels, and the 1000 Genomes Phase I gold standard indels as reference variants. The recalibration adjustment was then applied with ApplyBQSR.

### Variant calling

Deduplicated and recalibrated Hi-C reads were then processed using the GATK (v4.0.12.0) germline short-read variant discovery pipeline. Briefly, HaplotypeCaller was run in gVCF mode (flags “-ERC GVCF”) using dbSNP build 138 as a reference. Merged gVCFs then were converted to genomicsDB format with GenomicsDBImport and genotypes were called against this genomicsDB with GenotypeVCFs.

Variant quality scores were separately recalibrated for SNVs and indels via the GATK VQSR (variant quality score recalibration) procedure. Briefly, separate VQSR models were built for SNVs and indels using VariantRecalibrator, run in SNP or INDEL mode, respectively. The reference variants used for SNV quality recalibration were:

HapMap variants (v3.3): training and truth, prior of 15

1000 Genomes “Omni” platform variants (v2.5): training and truth, prior of 12

1000 Genomes Phase I gold standard SNPs: training only, prior of 10

dbSNP variants without 1000 Genomes (build 138, excluding sites after build 129): known, prior of 2

The reference variants used for indel quality recalibration were:

Mills + 1000 Genomes gold standard indels: training and truth, prior of 12

dbSNP variants without 1000 Genomes (build 138, excluding sites after build 129): known, prior of 2

The flags “--max-gaussians 2 -an QD -an MQ -an ReadPosRankSum -an FS -an SOR -an DP” were used when building the SNV recalibration model, and the flags “--max-gaussians 4 -an QD -an DP -an FS -an SOR -an ReadPosRankSum” were used when building the indel recalibration model.

The VQSR models for SNVs and indels were then applied using ApplyVQSR in SNP or INDEL mode, respectively, with a truth sensitivity filter level of 99.

### Haplotype phasing

Haplotypes were phased using HapCUT2. Briefly, recalibrated and filtered variants were separated for each sample, then HAIRS were extracted with extractHAIRS with flags “--hic 1 -- indels 1”. HAPCUT2 was then run with flag “--hic 1”.

Each Hi-C read was then assigned to one of the two haplotype blocks called by HapCUT2 by counting how many variants that overlapped the read were part of each haplotype block. If a read overlapped multiple variants that were phased to different haplotype blocks, a majority voting system was used to assign those reads to the haplotype block that had more variants overlapping that read. If an equal number of variants from each haplotype block overlapped the read, the read was discarded from the phasing process.

### Hi-C matrix construction and visualization

Hi-C matrices were constructed from mapped reads using the pairtools pipeline. Briefly, Hi-C read pairs were parsed, sorted, merged, and deduplicated. Restriction fragments were assigned to read pairs by using “pairtools restrict” with a restriction fragment bedfile generated using the “digest_genome.py” script from HiC-Pro.

Phased pairsfiles were generated by subsetting the unphased pairsfile to only those reads that were phased to a specific haplotype block.

Phased and unphased pairsfiles were used to assemble contact matrices using the “juicer pre” command in juicer_tools (v1.8.9), using a MAPQ threshold of 10. Phased matrices were assembled at 40 Kb resolution, while unphased matrices were assembled at 10 Kb resolution.

Unphased matrices were balanced using the KR (Knight-Ruiz) normalization implemented in juicer_tools and visualized in balanced form. Phased matrices were visualized in unbalanced form. H3K27ac ChIP-seq tracks from ENCODE (H1 neurons, Bernstein Lab, ENCODE ID ENCFF516KKW) were overlaid on the heatmaps.

## Supplemental Figures Legends

**Figure S1:**
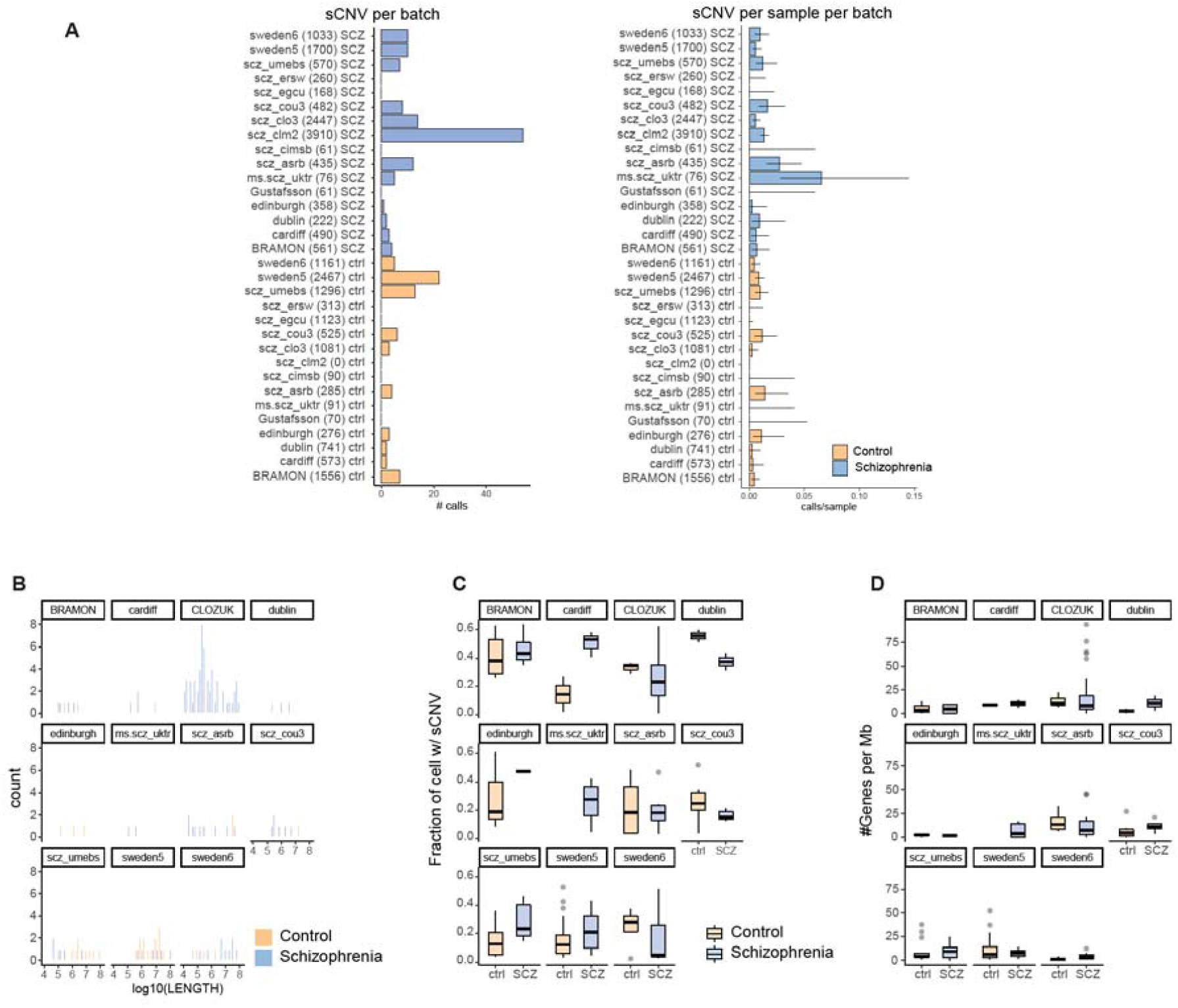
Characteristics of sCNVs callset across batches. A) Bar plots and forest plots of the number of sCNVs and fraction of samples with more than one sCNV in cases and controls for all batches of the data. The number of samples on each batch is indicated in the parenthesis of the y-axis labels. The 95% confidence intervals were calculated using the Wilson’s score interval with Newcombe modification. B) Histograms of sCNV length across batches for cases and controls. C) Box-plots of the fraction of cells with events (CF) in SCZ vs controls across all batches with events. D) Box-plots of the number of genes affected per megabase (Mb) in SCZ vs controls across all batches with events.

**Figure S2:**
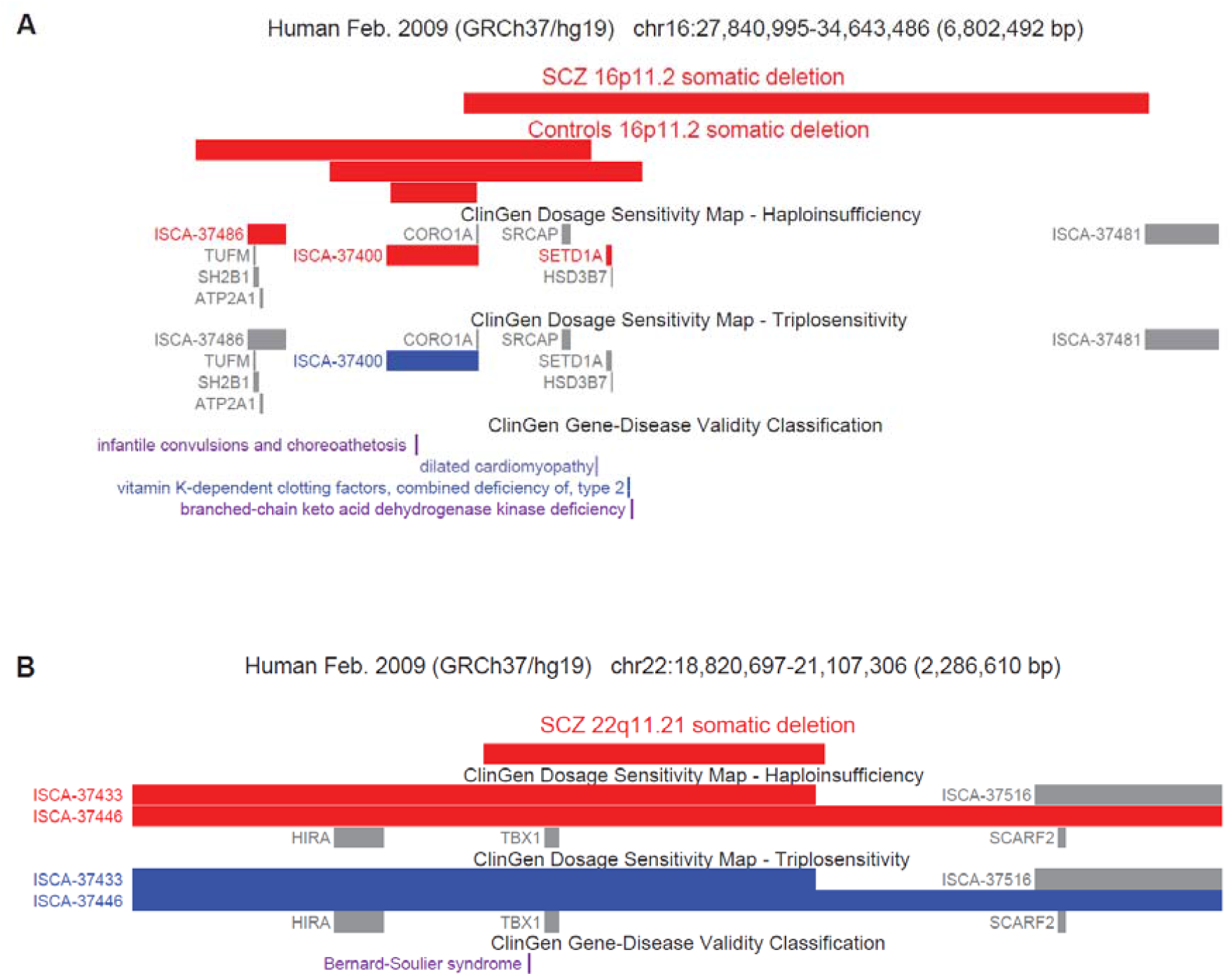
Somatic CNVs in 16p11.2 and 22q11.21. A) Adapted GenomeBrowser plot of 16p11.2 somatic deletions in cases and controls. Clinically relevant haploinsufficient and triplosensitive regions were annotated using the ClinGen database. Canonical 16p11.2 deletion regions are annotated by ClinGen haploinsufficiency at the proximal (ISCA-37400) and distal (ISCA-3786) sites. B) Adapted GenomeBrowser plot of 22q11.21 deletions in SCZ cases. The canonical 22q11.2 deletion regions are annotated as ISCA-37433 and ISCA-37446. For Figure A and B clinically relevant haploinsufficient and triplosensitive regions and genes were annotated using the ClinGen database. The red and blue color on in the dosage sensitivity map indicates deletions and duplications respectively. The gray color indicates that there is only moderate indication that the region/gene might be dosage sensitive. Note that *COMT* is overlapped by the 22q deletion, but is not illustrated because it is not part of the ClinGen annotation database.

**Figure S3:**
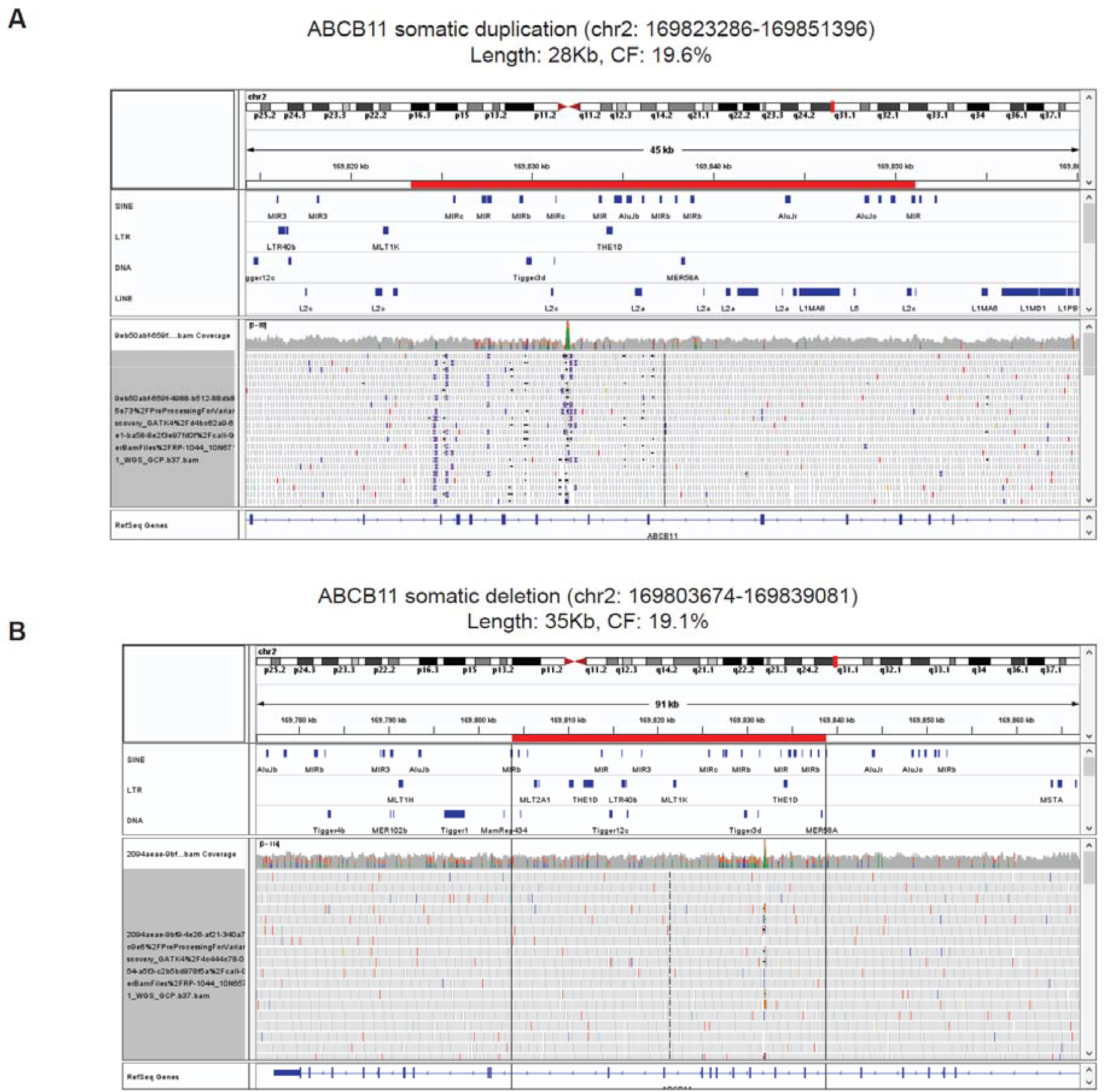
WGS IGV plots of *ABCB11* sCNV samples. A, B) IGV plots of *ABCB11* locus. Red bar representing the corresponding putative sCNV region. The tracks from top to bottom on each panel indicates the RepeatMasker annotation for different transposon families, coverage, and reads mapping to that region respectively.

**Figure S4:**
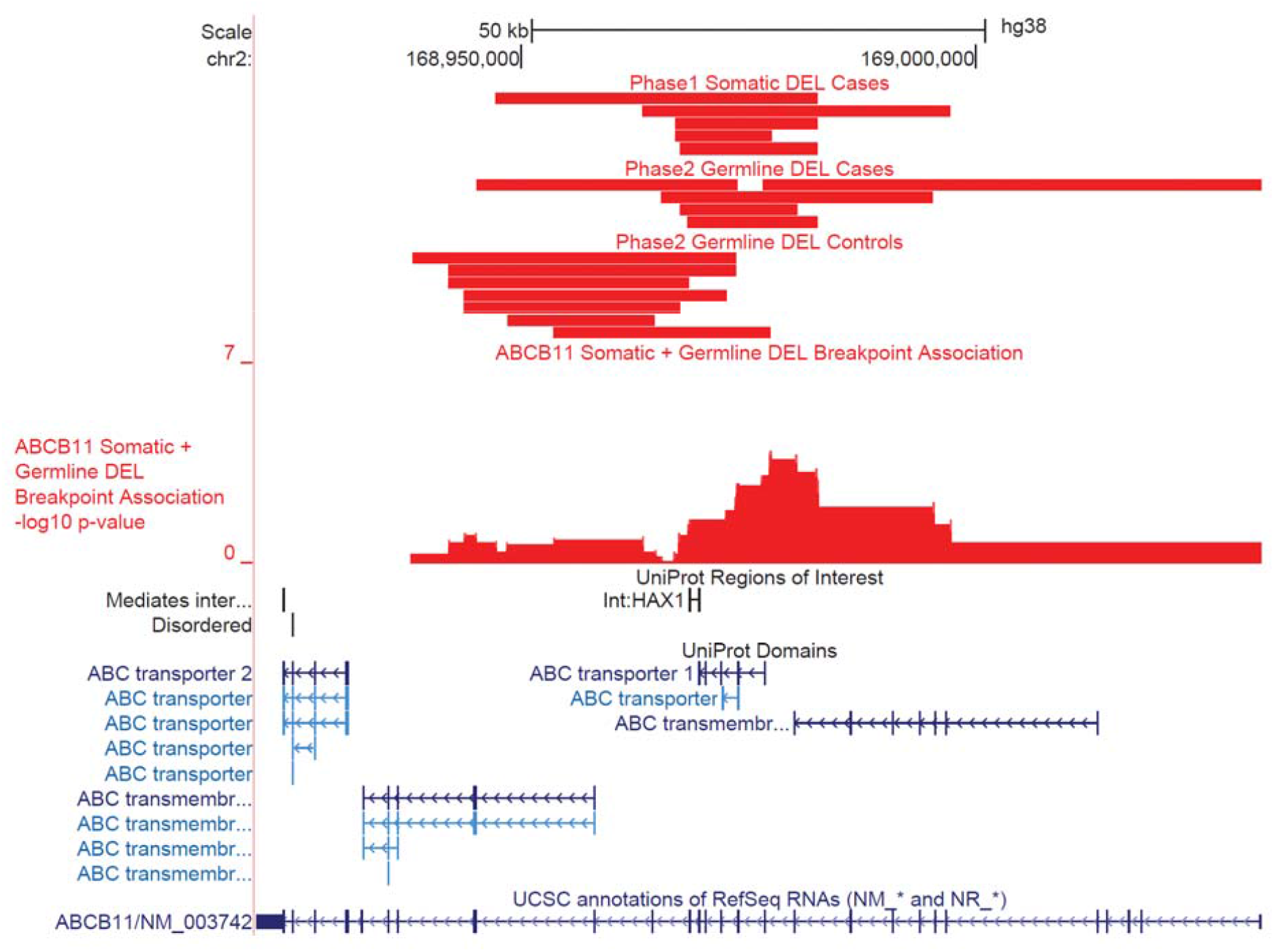
Somatic and Germline deletions of *ABCB11*. Adapted GenomeBrowser plot at the *ABCB11* gene locus. Association p-values were computed with logistic regression on disease status, controlling for overall CNV burden.

**Table S1: Membership and affiliations for Psychiatric Genomic Consortium and Brain Somatic Mosaicism Network**.

**Table S2: sCNV burden in SCZ cases and controls by gains and losses.**

**Table S3: Final sCNV callset of SCZ and Control samples**.

