## Supplementary figures and images for "Schizophrenia-associated somatic copy number variants from 12,834 cases reveal contribution to risk and recurrent, isoform-specific *NRXN1* disruptions"

### FigureS1

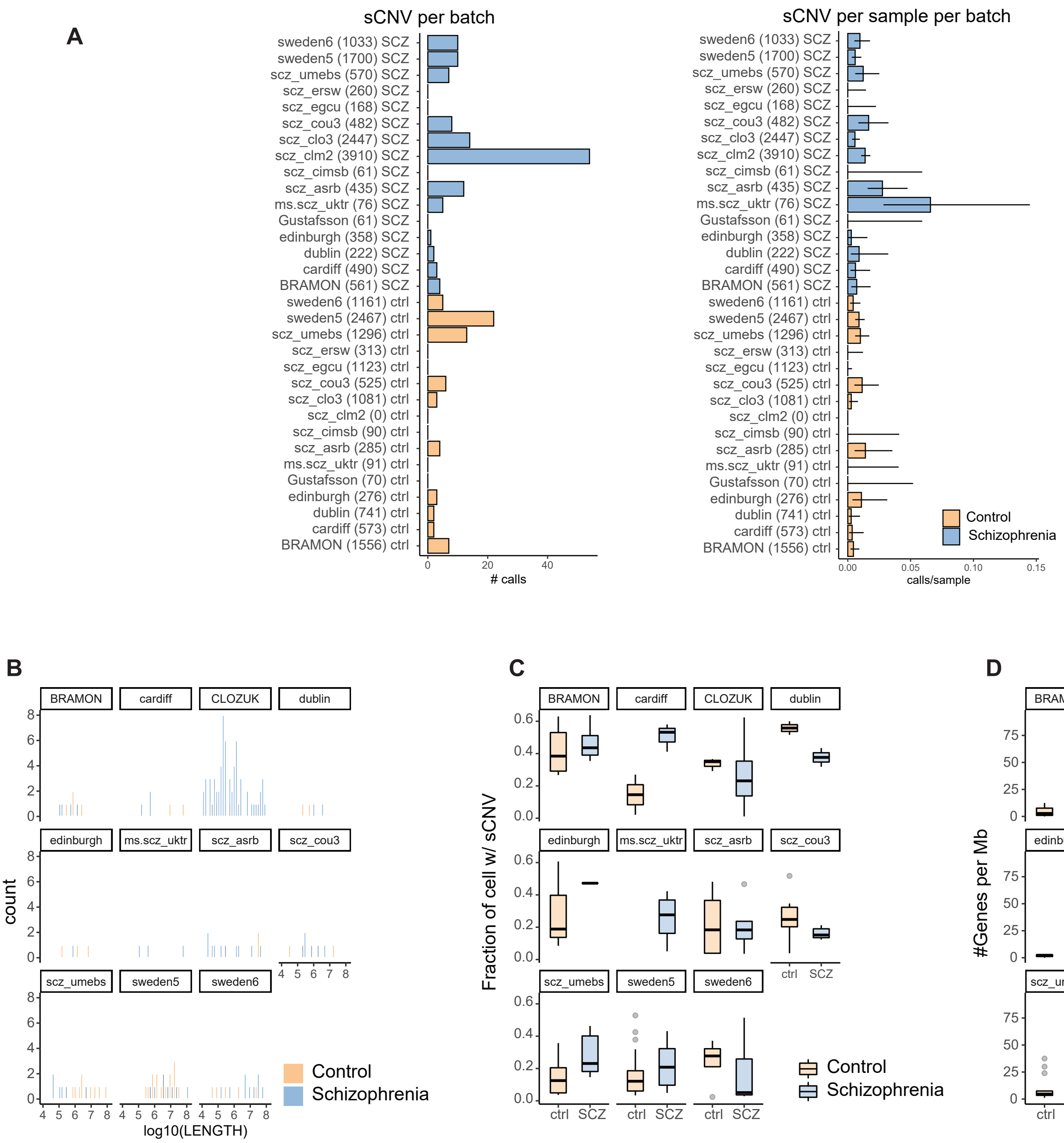

### FigureS2

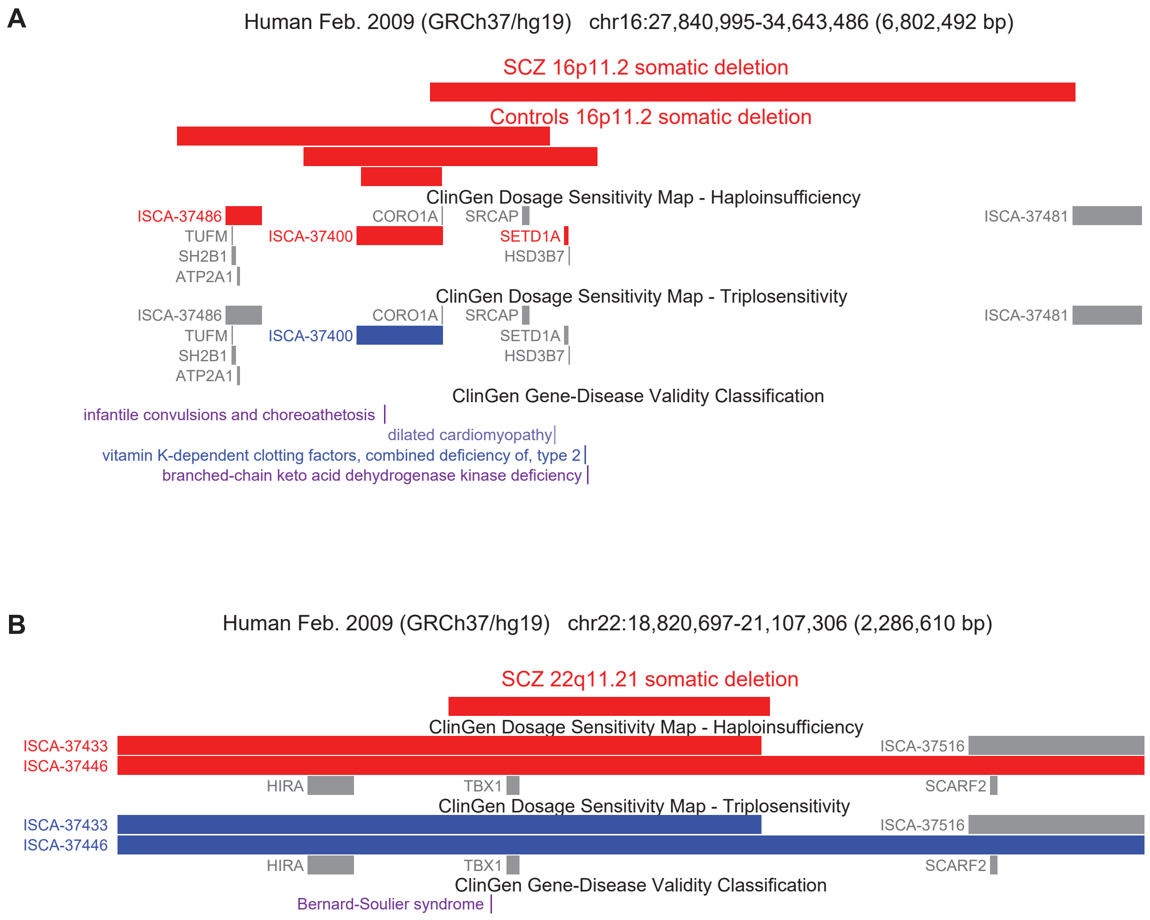

### FigureS3

A

ABCB11 somatic duplication (chr2: 169823286-169851396)
Length: 28Kb, CF: 19.6%

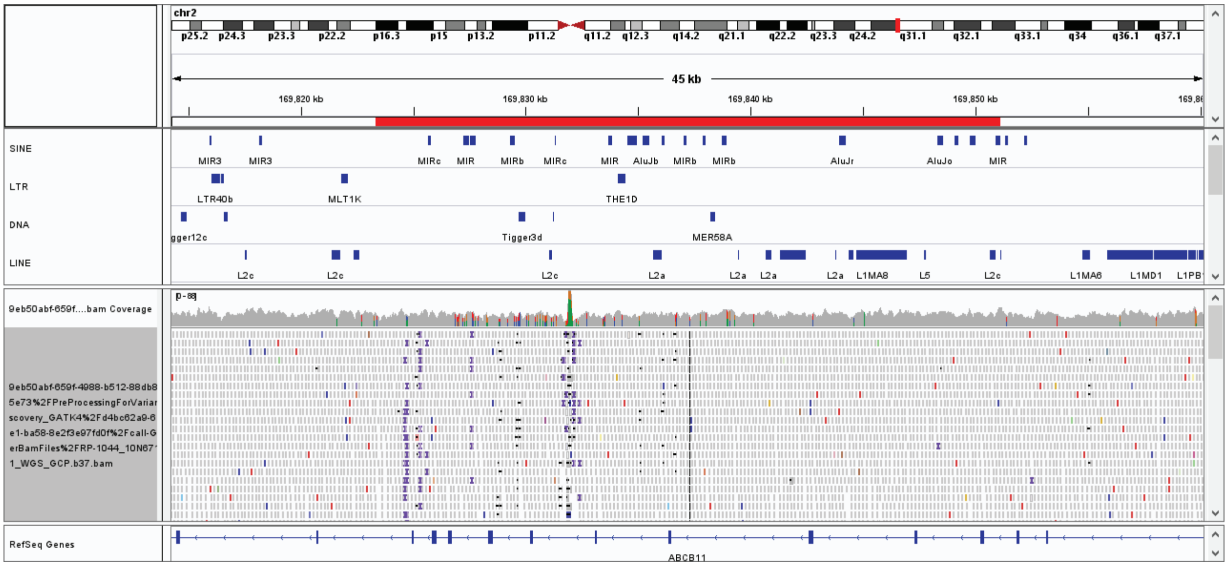


B

ABCB11 somatic deletion (chr2: 169803674-169839081)
Length: 35Kb, CF: 19.1%

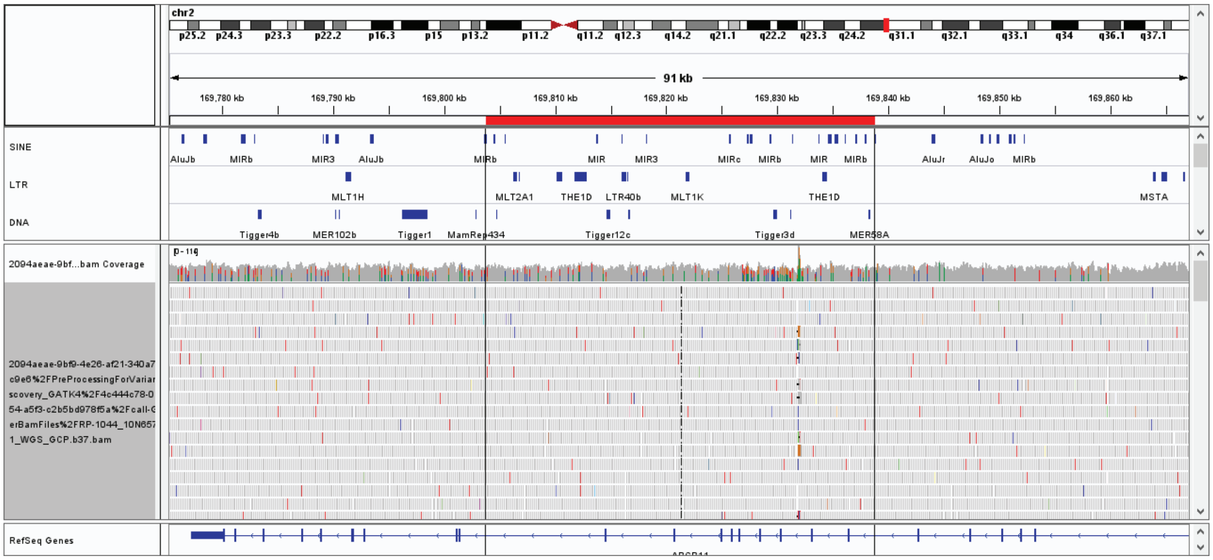

### FigureS4

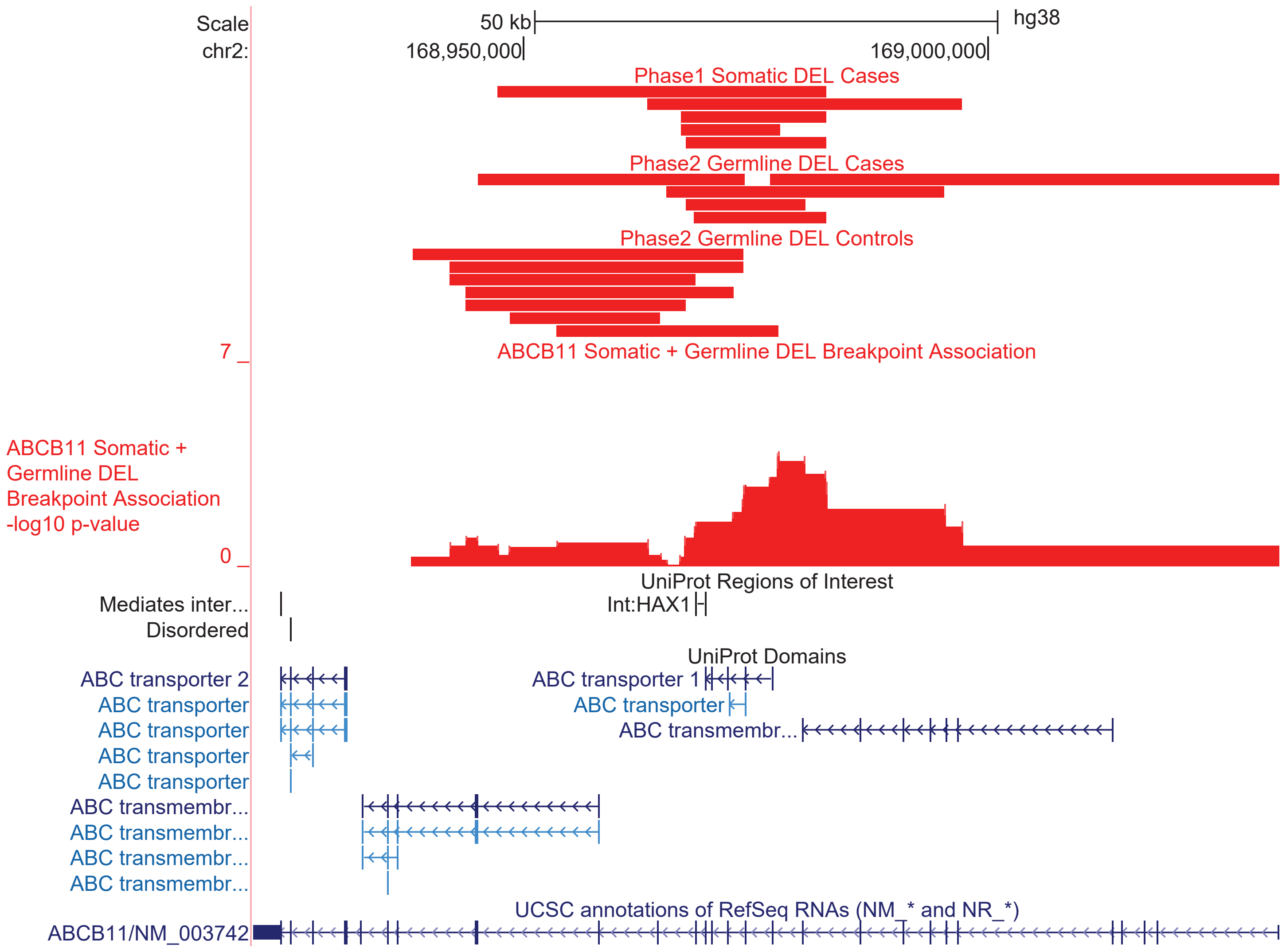
